# Characterization of LTBP2 mutation causing Mitral Valve Prolapse

**DOI:** 10.1101/2024.07.21.24302849

**Authors:** Shoshi Shpitzen, Haim Rosen, Ayal Ben-Zvi, Karen Meir, Galina Levin, Amichay Gudgold, Shifra Ben Dor, Rebecca Haffner, Donna R. Zwas, David Leibowitz, Susan Slaugenhaupt, Eyal Banin, Rotem Mizrachi, Alexey Obolensky, Robert A Levine, Dan Gilon, Eran Leitersdorf, Idit Tessler, Noga Reshef, Ronen Durst

## Abstract

**Background:** Mitral Valve Prolapse (MVP) is a prevalent valvular disorder linked to considerable morbidity and mortality, affecting approximately 2.4% of the general population. A prior genome association study linked LTBP2 to this trait. We report a knockout mouse with LTBP2 mutation demonstrating valve phenotype as well as a family with a novel mutation causing MVP

**Methods:** Exome sequencing and segregation analysis were conducted on a large pedigree to identify mutations associated with MVP. Using CRISPR-Cas9 technology, two strains of mice were generated: one with a complete knockout (KO) of the LTBP2 gene and another with a knock-in (KI) mutation corresponding to the putative causative mutation. Echocardiography and histological examinations of valves were performed in the KO and the KI at the age of 6 months. Optical coherence tomography (OCT) and histological examination of the eyes was done at the same time. mRNA qPCR analysis for TGFβ signaling targets (periostin/POSTN, RUNX2, and CTGF) in valve tissues was analysed.

**Results:** The LTBP2 rs117800773 V1506M mutation exhibited segregation with the MVP trait. LTBP2 KO mice had higher incidence of myxomatous changes by histology (7 of 9 of KO vs. 0 of 7 control animals, p=0.00186) and echocardiography (7 of 9 vs. 0 of 8, p=0.0011). LTBP2 Knock-in mice for the human mutation showed a significantly elevated myxomatous histological phenotype (8 of 8 vs. 0 of 9, p=0.00004) as well as by echocardiography (6 of 8 vs. 0 of 9, p=0.00123). KO mice demonstrated a significant increase in the depth of the anterior chamber as well as reduced visual acuity. LTBP2 KO mice demonstrated overexpression of both TGFβ signaling targets RUNX2 and periostin (P=0.0144 and P=0.001826, respectively).

**Conclusion:** Animal models of LTBP2 KO and KI recapitulate MVP phenotype indicating that LTBP2 mutations are indeed causing myxomatous degeneration. Further, LTBP2 rs117800773 V1506M segregated with MVP in a large pedigree. Our data indicate the importance of LTBP2 in normal mitral valve function and that mutations in the gene care causing myxomatous valve.

## Introduction

Mitral valve prolapse (MVP) is a very common cardiac valvular disorder that occurs in 2.4% of the general population ^1^. MVP is characterized by the displacement of one or both leaflets toward the left atrium during valvular closure during systole ^2–4^. Myxomatous alteration in the valvular tissue, changes in collagen organization and an increase in glycosaminoglycans, lead to biomechanically inferior valvular tissue that results in prolapse of the mitral leaflets into the left atrium ^3,4^. Prolapse of the leaflets may cause progressive degeneration and leakage, and therefore MVP is a leading indication for mitral valve surgery ^5^. MVP can be complicated by infective endocarditis, valvular regurgitation and congestive heart failure. In addition, several recent studies have demonstrated an association between MVP and ventricular arrhythmias and sudden cardiac death ^6,7^. Dysregulation of the extra-cellular matrix (ECM) components plays a key role in mediating these changes and is essential for understanding the genetic pathways causing the disease ^8^.

MVP is classified as non-syndromic or syndromic. Non-syndromic MVP can be familial or sporadic. Syndromic MVP occurs in association with connective tissue disorders such as Marfan syndrome, Loeys-Dietz syndrome, Ehler-Danlos syndrome, osteogenesis imperfecta, pseudoxanthoma elasticum and aneurysms-osteoarthritis syndrome. ^2^ Familial studies of idiopathic or non-syndromic MVP suggest an autosomal dominant model of inheritance with age dependent incomplete penetrance ^1–4^.

A prior genome wide association study suggested and association between LTBP2 (Latent Transforming Growth Factor Beta Binding Protein 2) and MVP. LTBP2 is a member of the larger TLBP family, proteins that are expressed in the ECM thought to have role in maintaining ECM functional and structural integrity. To confirm the causative role of LTBP2, we created two strains of mouse models, a LTBP2 Knock-Out (KO) model with a gene knock-out and a LTBP2 Knock-In (KI) model of the rs117800773 V1506M mutation that was found in a large pedigree segregating the trait. Both strains demonstrated high prevalence of MVP on echocardiogram and histology analyses.

## Material &Methods

### Genetic Analysis

The study was approved by the Hadassah Hebrew University Hospital in Jerusalem IRB and the Israeli Ministry of Health committee for genetic studies(0464-10-HMO). All participants consented to participate in the study. After signing an informed consent, subjects were clinically phenotyped by transthoracic echocardiography using a standard imaging protocol ^2,9^. MVP was defined by echo as leaflet displacement of at least 2 mm above the mitral annular line on the parasternal long-axis view (fig 1). DNA was extracted from peripheral blood leukocytes using the salting-out method. Exome sequencing was done on an Illumina platform, with a mean coverage of >150X. Bioinformatics was performed using the TGex NGS analysis platform ^10^. The Analysis platform was used for extraction of raw sequencing data from the sequencing provider, followed by primary and secondary pipelines to generate variant call format (VCF) files with exome variants. VCF files went through a comprehensive annotation pipeline for analysis, and all the resulting annotated variants were displayed in an interactive user interface for analysis interpretation and selection of plausible candidates in the context of MVP and related keywords. The annotation pipeline, as well as the annotation database and architecture of a variant interpretation of the platform, has been described in elsewhere in detail ^10^. Briefly, the analysis is based on the standard steps of basic variant annotation, allele frequency databases and variant damage prediction, and offers phenotype-driven interpretation that relies on a comprehensive knowledge base and annotation of structural variants. The genes were validated by PCR and Sanger sequencing.

**Figure 1:**
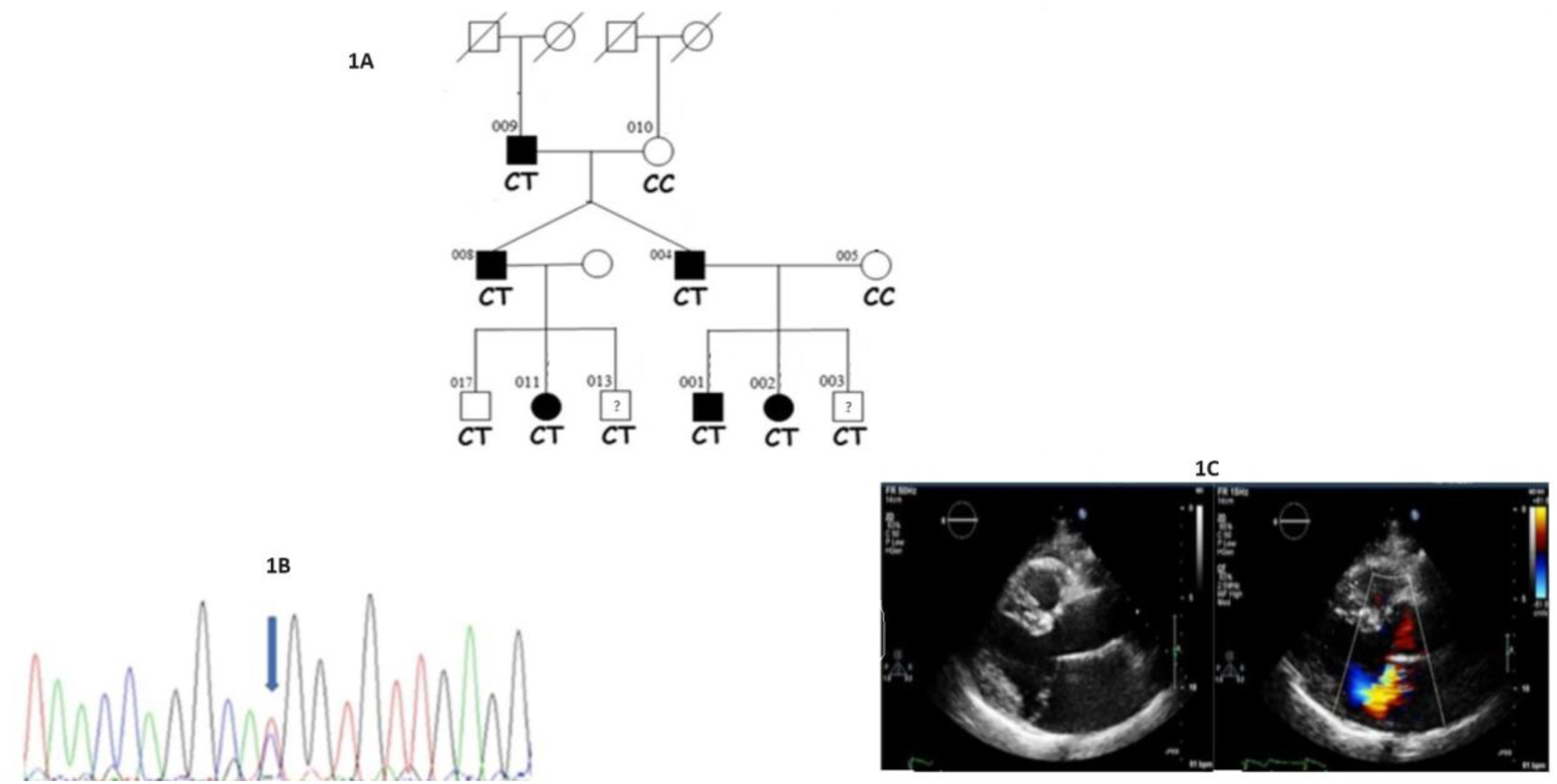
Pedigree demonstrating LTBP2 V1506M mutation. Segregation with MVP in part of the pedigree. The full pedigree is not disclosed to comply with MedrXives policies and regulation. Full disclosure of the pedigree is available upon request from the authors. Dark shapes are MVP, empty shapes are normal phenotype. A question mark denotes either equivocal phenotype or that an echocardiogram was not available. LTBP2 genotype is at the bottom: CC is the wild type allele and CT heterozygote for the V1506M mutation. Left top number is a serial number for the individual in the family. Fig1 B, the sequence variation with a red arrow pointing to the mutation. Fig1 C, a demonstrative parasternal long axis with and without color Doppler of individual 004, demonstrating MVP and significant regurgitation.

### Animal model

#### gRNA Design and the Generation of LTBP2 KO/I Mice

All mice experiments were approved by the IACUC committees of the Weizmann Institute and the Hebrew University Medical Center (MD-21-16445-3), and were carried out in accordance with their approved guidelines. gRNAs were designed using a combination of in-silico design tools, including the MIT CRISPR design tool ^11^ and sgRNA Designer, Rule set 2 ^12^, in the Benchling implementations ^13^ SSC^14^ and CRISPOR ^15^.

Two gRNAs were designed to target the LTBP2 region, LTBP2 upstream guide (5’-TTT GGC ACC GAG AAG CGA GC −3’) and LTBP2 downstream guide (5’-AAT CGG TTG TGG CAC CCC GT −3’) respectively, to create genetic KO of LTBP2.The planned deletion of 551 bp removes the beginning of the gene, including part of the promoter, the transcription start site, and the initiator methionine. We also designed a guide (5’-CGA TGC CTC AAC ATA GTG CC −3’) and an ssODN repair template (TCCCGAAGATGCCGATGAATGTGTACTGTTTGGGCCTGCTCTCTGCCAGAATGGCCGATGTCTGAATATTATGCCTGGATACATATGCCTGTGCAACCCTGGCTACC ACTATGATGCCTCCAGCAGGAAGTGCCAGGGTGAGGAC) to phenocopy the human mutation rs117800773 V1506M, corresponding to V1471M in the mouse (supplement figure 2). Cas9 nuclease, crRNA, tracrRNAs and ssODN were purchased from Integrated DNA Technologies (IDT). LTBP2 mice were generated at the Transgenic Facility at the Weizmann Institute of Science using CRISPR/Cas9 genome editing in isolated one-cell mouse embryos as described ^16^.

C57Bl/6JOlaHsd mice were purchased from Envigo, Israel and maintained in specific pathogen-free (SPF) conditions. Mice were maintained on a 12-hr light/dark cycle, and food and water were provided ad libitum. Cas9-gRNA ribonucleoproteins (RNP) complexes were delivered to one-cell embryos via electroporation, using the BioRad Genepulser (Bio-Rad, Hercules, California, USA). Electroporated embryos were transferred into the oviducts of pseudo pregnant ICR females (Envigo, Israel).

Genomic DNA from F0 pups was analyzed at weaning for the 551bp deletion, and theV1506M mutation by PCR and Sanger sequencing. The pups were genotyped by PCR (PCRbio HS Taq mix) and Sanger sequencing.

The validation of KO and the KI were done by using PCR with the following primers:

LTBP2 (KO) F 5’-GTT TGC CCA GGA CAG GAA AA −3’

LTBP2 (KO) R 5’-GTT CCA CTC AGA GGG CGA −3’

LTBP2 3F (KI) 5’-TCC TCC CGA AGA TGC CGA −3’

LTBP2 3R (KI) 5’-CGT TCT CAC AGG CCA AGT CC −3’

#### Western blot analysis

To validate reduction in LTBP2 protein in the knockout mice, total protein was purified from the whole heart using homogenization in T-PER buffer (tissue protein extraction reagent Thermo scientific) and protease inhibitor (Pierce protease inhibitor –tablets A32953 Thermo scientific) analyzed by 4-12% SDS-PAGE (Thermo fisher scientific) and immunoblotting with LTBP2 antibody 1:100 (LTBP2 (E-10) SC-166199), and goat anti mouse as a secondary antibody 1:10000 (Jackson immunoresearch laboratories cat:115035166) (supplement figure 1).

#### Animal Echocardiography

Echocardiography was performed at 6 months of age. Both the pathologist and the cardiologist performing the echocardiography were blinded to the underlying genotype. Echocardiography was performed using the Vevo3100 Lazer-x Ultrasound (Visual Sonic Ltd, London, UK) at The Wohl Institute for Translational Medicine at the Hadassah Hebrew University Medical Center. After sedation with isoflurane, the animals were placed in prone position. A parasternal long axis view was obtained. Displacement of the posterior leaflet was confirmed if the leaflet traversed the annular line drawn between the hinge points of anterior and posterior leaflets. Other parameters that are related to MVP, such as anterior displacement of the coaptation line were used to confirm the phenotype ^17,18^.

#### Histological analysis

LTBP2 knock-out and knock-in mice were sacrificed for histopathologic analysis at 6 months. Mitral valve disease progression was examined using the murine model of both strains of LTBP2. For histopathological analysis hearts were processed in 4% formaldehyde. Paraffin embedded tissue slices included left ventricular apex, mitral valve, and the aortic valve. Thin slices of the hearts were prepared using microtome and placed for H&E staining and Movat Pentachrome staining. Movat’s stain (Movat pentachrome stain kit ab245884) is a pentachrome stain that highlights the various constituents of connective tissue and H&E. It differentiates collagen, elastic, muscle, mucin, and fibrin by color (muscle in red, glycosaminoglycan blue, elastin dark purple, nuclei in black and collagen yellow.)

#### Immunofluorescence staining for LTBP2 deficient valve tissue

LTBP2 KO and WT mice were sacrificed at 6 months by cervical dislocation. Hearts were processed in 4% formaldehyde. Paraffin embedded tissue sectioned at 4-μm thickness included left ventricular apex and mitral valve. Slides were deparaffinized in xylene and dehydrated through a graded ethanol series. The slides were then transferred to a pressure cooker in 10 mM citrate buffer ph6.0 (Antigen Unmasking Solution, citric acid-based vector H-3300) for antigen retrieval. Blocking section were done in CaseBlock 10 (Abcam AB64226) for 10 min and incubated with LTBP2 antibodies 1:200 (kind gift of Dr. Gerthad Sengle from the Center for Molecular Medicine Cologne Germany) overnight in 4^0^C. Alexa flour conjugated 647 Donkey anti rabbit 1:200 (Jackson Immunoresearch 711605152) was used as a secondary antibody for 45 min at room temperature. The section was mounted with DAPI mounting (DAPI Fluoromount-G SouthernBiotech 0100-20) and covered with coverslip. The sections were observed and images were taken with a confocal fluorescence microscope Nikon Spinning Disk Microscope (Yokogawa W1 Spinning Disk. 2 SCMOS ZYLA cameras x40 objective 200 msc exposure). The processing was done by NIS Elements software package for multidimensional experiments, with JOBS for high content acquisition and analysis, deconvolution, tracking,3D automatic measurements.

#### qPCR

After sacrifice hearts were preserved on ice cold saline solution. The hearts were dissected under a binocular from the aorta to the apex. Mitral valve tissue was identified and excised. Total RNA was extracted from heart valve using Tri-reagent (sigma T9424) according to manufacturer’s protocol, For reverse transcription reaction we used Quanta bio kit (qScript cDNA synthesis kit 95047-100 Quantabio, Beverly, MA, USA) and q-RT-PCR was done on 3 genes: mouse RUNX2, mouse Periostin and mouse CTGF using Syber green fast mix (Quanta bio 66185483). Rodent GAPDH was used as standard. Primers for the reactions were: RUNX2 F 5’-AGA GCC AGG CAG GTG CTT C-3’, RUNX2 R 5’-TCT CAG TGA GGG ATG AAA TGC T −3’,

Periostin F 5’-CCA CAG GAG GTG GAG AAA CA −3’, Periostin R 5’-GAT CGT CTT CTA GGC CCT TGA-3’,

CTGF F 5’-GGA GTG TGC ACT GCC AAA GA −3’, CTGF R 5’-ACT CAC CGC TGC GGT ACA C-3’ The samples were analyzed on ABI-7300 Real Time PCR system (Applied Biosystems, Waltham, MA, USA)

### Mice eye analysis

#### Visual acuity (VA)

VA of wild-type (WT) and LTBP2 knockout (LTBP2 KO) mice was evaluated using the optomotor response test (OMT) (OptoMotry; Cerebral Mechanics, Lethbridge, AB, Canada), as previously described^19^. Animals were placed on a small platform at the center of a virtual drum presented by four liquid crystal display (LCD) panels. After a short adaptation period, VA was measured at 100% contrast by recording the tracking response to a rotating visual stimulus.

#### In vivo imaging

Anterior chamber structure was studied in vivo using single horizontal 15⁰ Optical Coherence Tomography (OCT) b-scans passing through the center of the cornea (SPECTRALIS, Heidelberg). The procedures were performed in anesthetized animals (intraperitoneal injections of a mixture of 0.85 μl Ketamine (Bedford Laboratories, Bedford, OH) and 0.15 μl Xylazine (VMD, Arendonk, Belgium) with dilated pupils (cyclopentolate 1% applied twice, Laboratorio Edol, Carnaxide, Portugal and tropicamide 1% applied once, Fisher Pharmaceuticals, Tel-Aviv, Israel). Anterior chamber depth (ACD, between corneal endothelium and anterior capsule of the lens), iridocorneal angle and pupil diameter were measured using Image Pro Plus 6.0 software.

#### Eye Histology

Eyes were enucleated, fixed in Davidson solution, embedded in Paraplast, and sectioned at 5 μm thickness through the center of the optic nerve. For descriptive histology, sections were stained with hematoxylin and eosin. All observations, photography and processing were performed using Apperio CS2 slide scanner and ImageScope software.

### Statistical analyses

Comparison between categorical parameters was made by chi square test. P values of each qPCR experiment was repeated three times for each set of RNA mitral valve, and results were pooled for statistical analysis. Student’s two tailed T-test was used for comparison between two groups. Movat’s pentachrome and Echocardiogram analyzes was made by chi square test. Eye imaging parameters were tested using a two-tailed Student’s t-test.

## Results

We studied 4 generations of a family comprising 28 individuals. The family had a severe clinical phenotype of MVP as several developed severe valvular disease requiring valve repair surgery (Figure 1). The family lacks extra cardiac connective tissue features and all do not fulfil any of the criteria for connective tissue disorders ^20–22^.The severity of mitral regurgitation (MR) was more noticeable among middle-aged individuals within the family. Three middle aged individuals had valve surgery (#004, 008, and 009). In contrast, younger members displayed a milder form of MVP with non-significant MR. Four family members (#004,008,009,011) were screened for a shared mutation by exome sequencing. 361 variants in 279 genes were found. Among these, pathogenic autosomal dominant mutations were identified in 11 genes (table 1). These were tested for segregation with the trait in the entire family.

**Table1:**
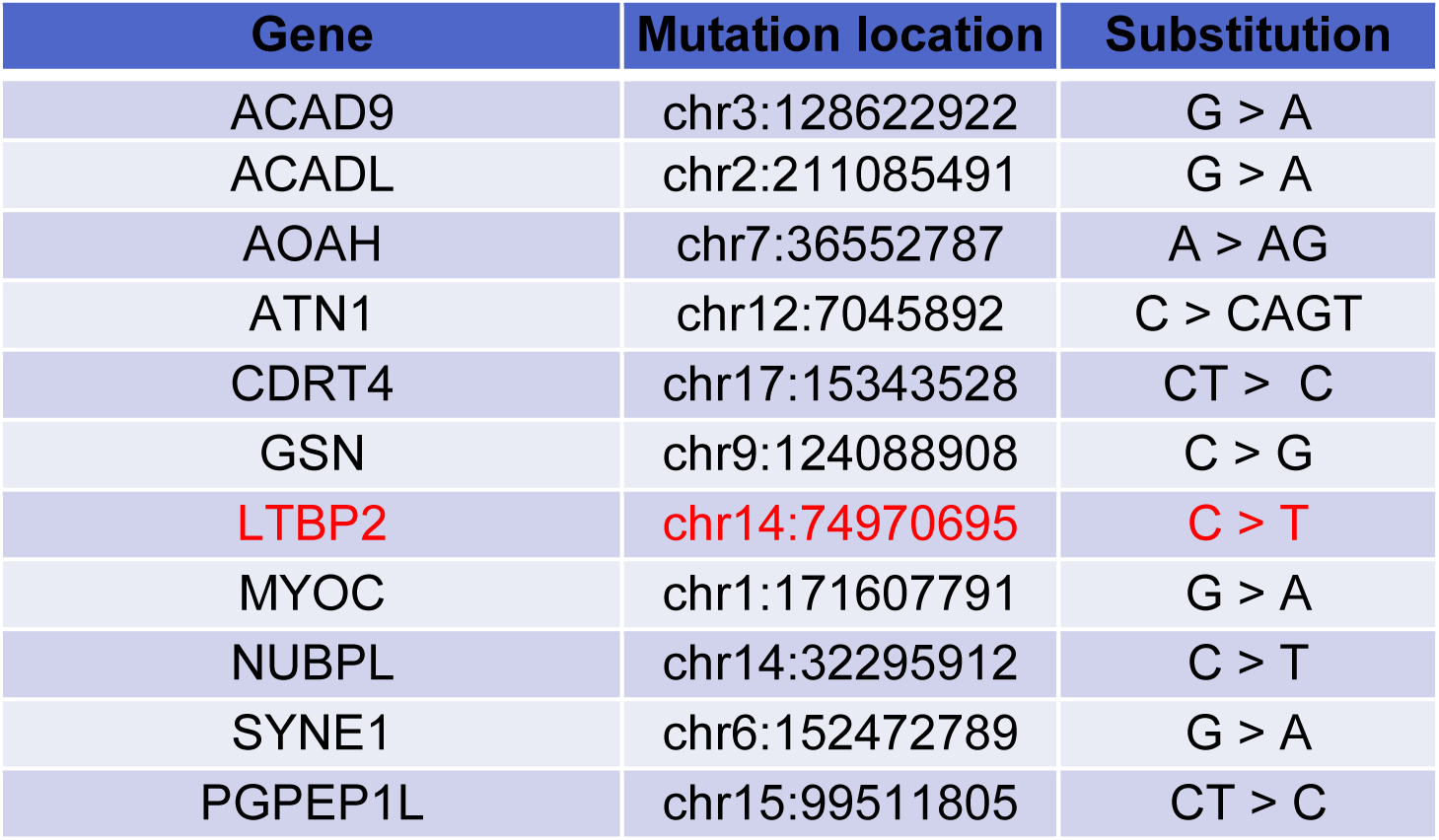
Next-generation sequencing analysis was performed on 4 affected family members and reveals all the genes that have been associated with the clinical data and reasonable functional influence on the protein structure. All the family members were tested for segregation and only LTBP2 V1506M mutation segregated with MVP in the extended family pedigree.

| Gene | Mutation location | Substitution |
| --- | --- | --- |
| ACAD9 | chr3:128622922 | G > A |
| ACADL | chr2:211085491 | G > A |
| AOAH | chr7:36552787 | A > AG |
| ATN1 | chr12:7045892 | C > CAGT |
| CDRT4 | chr17:15343528 | CT > C |
| GSN | chr9:124088908 | C > G |
| **LTBP2** | **chr14:74970695** | **C > T** |
| MYOC | chr1:171607791 | G > A |
| NUBPL | chr14:32295912 | C > T |
| SYNE1 | chr6:152472789 | G > A |
| PGPEP1L | chr15:99511805 | CT > C |

Only the LTBP2 rs117800773 V1506M (chr14:74970695) mutation showed segregation with trait. (Fig 1). rs117800773 V1506M is a rare mutation (minor allele frequency of 0.0001). The LTBP2 gene was a leading candidate as it was also found to be positively associated with MVP on a recent large GWAS (7), supporting our finding. In addition, LTBP2 is associated with TGFβ signaling activity, which has been shown to be disrupted in mitral valvulopathy in the setting of Marfan’s syndrome ^23^.

As these data supported the hypothesis that an LTBP2 mutation led to myxomatous degeneration in both a family and a GWAS study, the next step was to investigate whether LTBP2 mutation in this gene can cause myxomatous degeneration in mice. We therefore established two lines of mice using CRISPR –Cas9 technology, one with a major deletion (551bp) at the 5’ end of the gene, including part of the promoter, the transcription start site, and the initiator methionine resulting in complete knockout of the gene and the other, a KI of the rs117800773 V1506M mutation strain. The mouse strains were verified by PCR followed by Sanger sequencing and by Western-blot analysis, using an LTBP2 specific antibody (see supplementary figures 1 and 2).

### Exploring the KO and KI phenotypes by both functional imaging (Echocardiogram) and structural by histological examination (figure 2 and table 2)

Whole heart extracts were used for Western blot analysis of LTBP2. KO animals lacked LTBP2 protein expression in heart tissue (supplement figure 1). The leaflets of KO mice did not exhibit LTBP2 in immunofluorescence staining (Figure 2 G, H)

**Fig 2:**
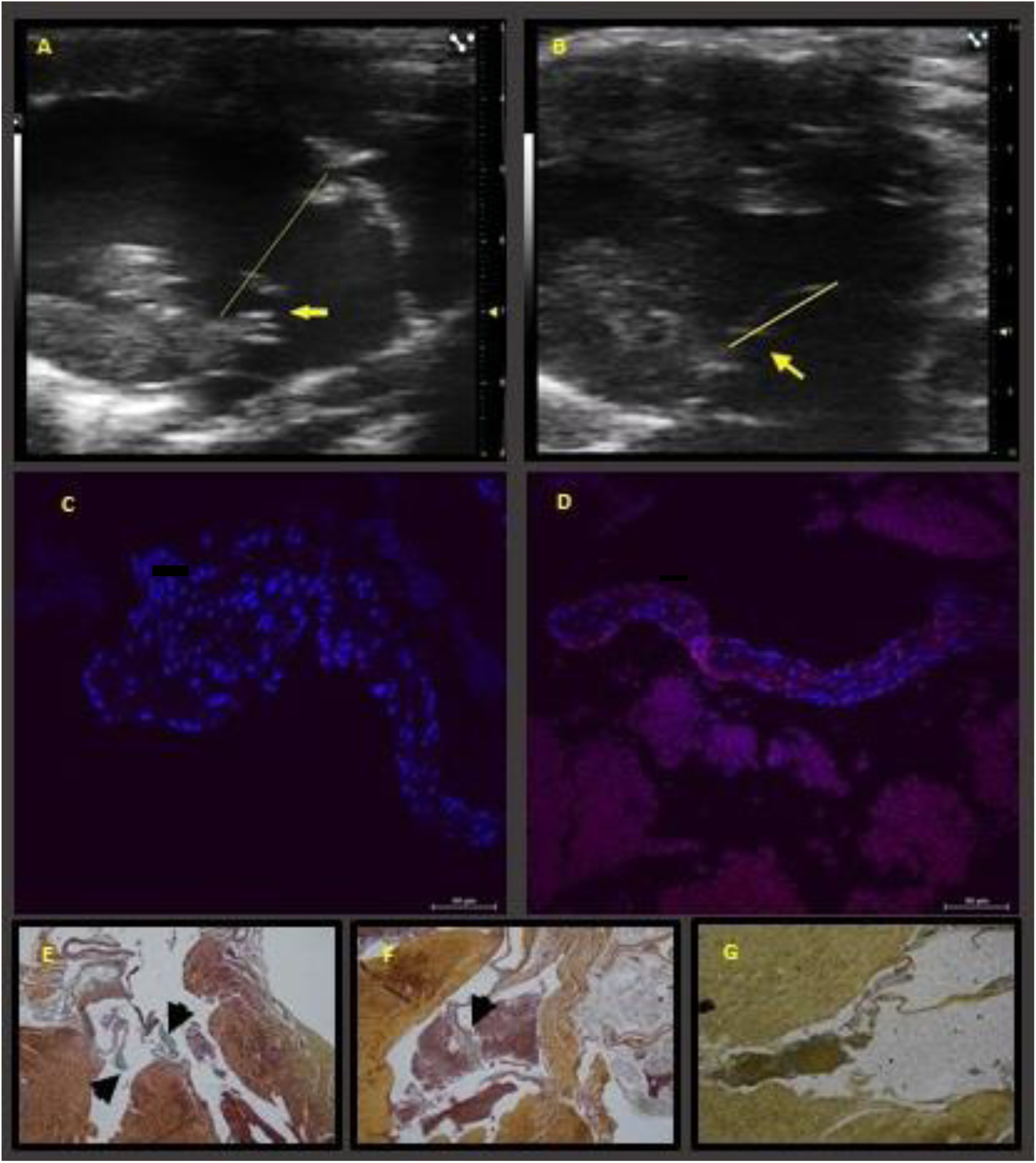
Valve phenotype of LTBP2 deficiency: echo cardiograms of 6 months old KO (A) and wild type (B). The yellow line delineates annular line. The arrow is pointing to the posterior leaflet. Mitral valve LTBP2 immunofluorescence in a KO mouse (C) and wild type (D). Notice the Cy5 coloring (red) of the wild type valve that is lacking in the KO. E, F and G are Movat pentachrome straining of KO, KI and wild type mice respectively. Notice the marked thickening and fibrosis with myxomatous changes of the valve leaflets in the KO (black arrows) (E) and the widespread blue staining of mucinous substance (arrow) (F). The leaflets of the wild type mice are thin and are not stained blue (G).

**Table 2:**
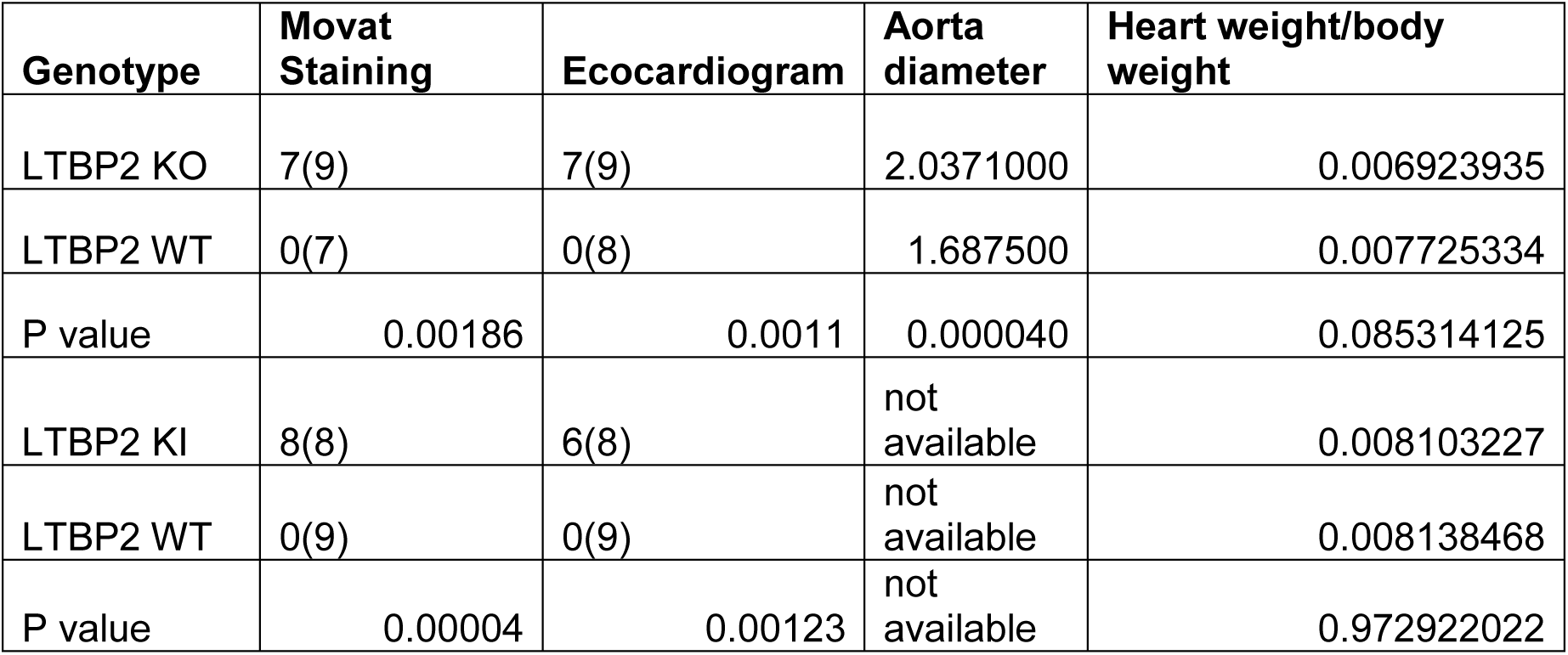
Represents the number of positive phenotypes among wild type (WT) and mutated animals for myxomatous degeneration by histology, valve and echocardiography. The absolute number of animals, out of total (n) and the percent on the bottom line. P value is calculated by chi test. KO=complete LTBP2 knockout. KI homozygous for LTBP2 V1506M mutation

#### Histology

Movat’s pentachrome staining and H&E demonstrated structural expansion of the valvular extracellular matrix (ECM), as well as thickening and myxomatous degeneration (fig 2). KO mice demonstrated a significantly higher rate of myxomatous changes by histology [7 of 9 of the affected mice vs 0 of 7 control animals (p<0.00186)]. KI mice demonstrated with significantly higher phenotype rate by histology [8 of 8 vs. 0 of 9 (p<0.00004)]. (figures 2 A-D and table 2).

#### Echocardiogram

Seven out of 9 KO animals showed leaflet prolapse compared to none of the 8 in the control group (p=0.0011). KI mice demonstrated a higher incidence of the phenotype as well, with 6 out of 8 exhibiting it compared to none of 9 in the control group (p=0.00123) (refer to figures 2 E, F).

#### qPCR for RUNX2 and periostin

Ample of evidence suggests that the TGF β – LTBPs – fibrillin complex is important for both ECM integrity and appropriate TGFβ/cytokine signaling. Given that LTBPs are regulators of TGFβ signaling we analyzed the expression of three TGFβ target molecules in valve tissue RNA extracts: RUNX2 (Runx family transcription factor 2), Periostin and CTGF (a member of the connective tissue growth factor family) ^24^. Significant overexpression of both Runx2 and Periostin was found in LTBP2 knockout valve tissue (RUNX2 P=0.0144 and Periostin P=0.001826). There was no significant difference in CTGF expression.

### Exploring Eye phenotype

*In vivo* OCT imaging of the anterior chamber of the eye demonstrated a statistically significant increase in the depth of the anterior chamber in 12-14 months old *LTBP2* KO mice as compared to age-matched control mice (746.3±27.1 µm versus 376.3±4.6 µm in WT eyes, mean±SEM, p<0.0001; Figure 3 A). Additionally, a substantial increase in the iridocorneal angle of the anterior chamber was identified (50.5±1.2 degrees versus 35.6±1.1 degrees in WT eyes, p<0.0001; Figure 3 B).

**Figure 3.**
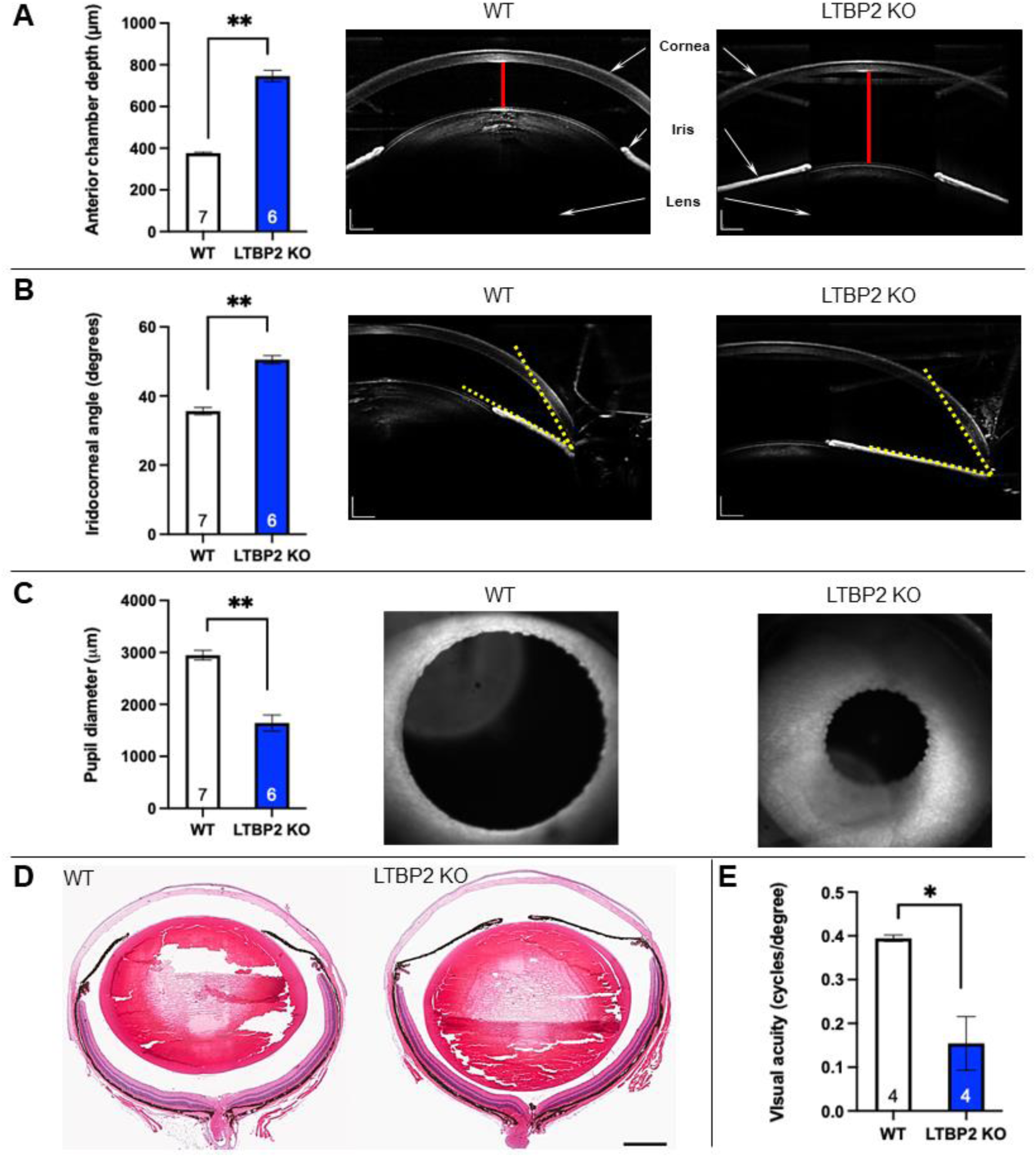
Ocular structure and visual function in 12-14 months old *LTBP2* KO mice. White bars represent WT mice and blue bars represent *LTBP2* KO mice. Results are presented as mean ± SEM. Number of animals in each group is written within the corresponding bar. * - p<0.05, ** - p<0.0001. **A.** Comparison between WT and *LTBP2* KO mouse eyes revealed a significant increase in the anterior chamber depth (ACD) in mutant mice as measured using *in vivo* OCT imaging of the anterior segment. Representative images from 12 months old mice are shown to the right of the bar graph. ACD reflects the distance between the corneal endothelium and the anterior capsule of the lens and is marked by red lines. Scale bars = 240 micrometers. **B.** Knockout of the *LTBP2* gene also led to significant enlargement of the iridocorneal angle. Representative images of iridocorneal angle (marked by yellow dotted lines) in 12 months WT and *LTBP2* KO mice. **C.** From 12 months of age and on, *LTBP2* KO mice exhibited very poor dilation of the pupils in response to topical application of mydriatic drops. Measurements revealed a statistically significant reduction in pharmacologically dilated pupil diameter in mutant mice compared to age-matched WT mice. Representative images of eyes of 12 months old mice captured in infrared mode are presented to the right of the graph. **D.** Representative images of eye sections from 12 months old mice, taken through the central cornea and optic nerve and stained with hematoxylin and eosin, align with our *in vivo* optical coherence tomography (OCT) findings. Mutant eyes show a deeper anterior chamber caused by posterior subluxation of the lens accompanied by an increased iridocorneal angle, reduced depth of the vitreous cavity, and a narrow pupil. Scale bar = 500 micrometers. **E.** Visual acuity as measured using the optomotor response revealed a significant reduction in *LTBP2 KO* mice compared to WT mice.

Furthermore, from 12 months of age and on, *LTBP2* KO mice exhibited very poor dilation of the pupils in response to topical application of mydriatic drops.

Measurements revealed a statistically significant reduction in pupil diameter following application of cyclopentolate and tropicamide dilating drops in mutant mice compared to age-matched control mice (1641.8±153.1 µm versus 2946.3±90.5 µm in WT eyes, p<0.0001; Figure 3 C). The results obtained by *in vivo* OCT imaging correlated well with *ex vivo* histological findings of the eye (Figure 3 D). In addition, histological sections showed that in the eyes of mutant animals, there is displacement of the lens towards the posterior pole of the eye, significantly reducing vitreous chamber depth and bringing the lens in close approximation to the retina. The structural changes led to impairment of visual function. Visual acuity as measured using the optomotor response in adult (12-14 months-old) mutant animals revealed a dramatic reduction in acuity compared to age-matched control mice (0.15±0.1 cycles/degree versus 0.39±0.0 cycles/degree in WT eyes, p<0.05; Figure 3 E).

## Discussion

Recently, Roselli et al described an association between LTBP2 and MVP in a large GWAS ^25^. We identified a family with MVP with an LTBP2 rs117800773 V1506M mutation that segregated with the trait with a high degree of penetrance (10 individuals out of 28). we created two mouse models, the LTBP2 KO and the LTBP2 KI carrying the rs117800773 V1506M mutation. Both strains exhibited a high prevalence of MVP, as observed through echocardiography and histological analysis. Moreover, the KO animals displayed a significant aortic dilation. Furthermore, in contrast to WT mice, mutant mice lacking LTBP2 exhibited posterior displacement of the lens leading to a significant increase in anterior chamber depth and a wider iridocorneal angle. Our combined findings from both human and animal models substantiate the involvement of LTBP2 mutations in the development and progression of MVP, elucidating their potential role in the pathogenesis of the condition. Our KO model manifests a pronounced impact on myxomatous degeneration, underscoring the significance of this protein in both valve function and structure. As expected and demonstrated before, LTBP2 deficiency results in eye anterior chamber phenotype in mice. The KI effect suggests that the identified human mutation is pathogenic. However, the mutation may also be in linkage disequilibrium with another mutation in the gene in this family. Interestingly, not all mutated mice exhibited a mitral valve prolapse (MVP) similar to humans, where penetrance is incomplete. This observation suggests that MVP is a progressive phenomenon and penetrance may be age-dependent ^17^.

We also describe a large family with non syndromic isolated type of MVP. Some of the family members have severe form of the disease. It seems that the disease severity is age dependent as MR severity as well as the need for surgical repair appears only in middle aged individuals. Age dependent progression of MVP phenotype was demonstrated before by Delling in the Framingham heart cohort ^17^. The significant association of LTBP2 with MVP in two distinct cohorts and using two different genetic approaches, strongly supports its role as a causative gene for the disease.

The LTBP2 gene, mapped to chromosome 14, is an isoform of the LTBP superfamily of extracellular matrix proteins. These are large, secreted glycoproteins structurally related to fibrillins. LTBP2 is expressed abundantly and its expression is particularly strong in tissues enriched in microfibrils, such the aorta, lung, heart, thyroid, ovary and testis ^26,27^. LTBP2 is associated with ECM proteins, TGFβ, fibrillin, and elastin regulation pathways. However, the specific roles of LTBP2 in ECM function is unknown. LTBP2 is a unique member of the family as it is the only isoform that does not directly bind to latent TGFβ ^28^. LTBP2 not only co-localizes with fibrillin microfibrils, but its deposition is dependent on preformed fibers of fibrillin-1. LTBP2 competes with LTBP1 to bind to the same binding site in fibrillin containing microfibrils, leading to the release of LTBP1 from microfibrils (19), and thus may indirectly negatively regulate the activation of TGFβ by releasing LTBP-1 from microfibrils. Indeed, deficiency of the long form of LTBP1 in mice causes serious disruption of great vessels and cardiac valve development, resulting in perinatal death ^29–31^. In our animal model, we demonstrated increased expression of TGFβ effector genes, RUNX2 and periostin. These play an important role in cardiac development and are known regulators of ECM remodeling. RUNX2 is a TGFβ and bone morphogenetic protein regulated gene required for epithelial mesenchymal transition. Periostin is a secreted fasciclin-domain-containing protein, which is involved in both valve development and valvular heart disease (30). Several studies demonstrated the importance of periostin in developing heart valves. Mice lacking periostin showed irregular matrix organization as periostin promotes cellular organization and differentiation of mesenchymal cells ^32,33^. RUNX2 is a regulator of endothelial mesenchymal transformation, a process important in normal valve development ^34^.

The importance of fibrillin for valve structure was elucidated in the Marfan Syndrome (MFS). Marfan syndrome (MFS; MIM 154700) is a relatively common autosomal dominant hereditary disorder of connective tissue with prominent manifestations in the skeletal, ocular, and cardiovascular systems. The changes typically seen in the cardiovascular system are dilatation of the aorta and MVP. MFS is caused by mutations in the gene for fibrillin-1 (FBN1)^35–37^. Fibrillin 1 assembles into microfibrils that serve a critical role in the maintenance of the structural integrity of the aortic wall, as well as the ciliary apparatus supporting the ocular lens. Hence, FBN1 mutations were initially thought to lead to tissue fragility, exclusively through disintegration and fragmentation of the connective tissue fibers. A revolutionary shift in thinking about the mechanisms of MFS occurred by studying lung disease in fibrillin 1-deficient mice. In the lungs of developing mice, increased levels of free TGF-β in addition to the downstream effectors of TGF-β signaling (pSMAD2/3) coincided with primary failure of distal alveolar septation. In vivo TGF-β antagonism using neutralizing anti-TGF-β antibody prevented the lung phenotype. Similarly, involvement of dysregulated TGF-β signaling in the etiology of MFS was subsequently established by Ng et.al. for the myopathy, mitral valve prolapse, as well as the aortic aneurysmal phenotype ^23^. As stated above, TGF-β interacts with fibrillin in the ECM through the LAP complex. Because of this interaction, TGF-β bioavailability is meticulously controlled by cytokine sequestration into the ECM LAP complex. Fibrillin 1 deficiency, due to FBN1 mutations, impairs ECM targeting of the LAP, resulting in an unrestrained release of TGF-β ligands ^37^. Evidence suggests that the TGF β – LTBPs – fibrillin complex is important for both ECM integrity and appropriate TGFβ/cytokine signaling. Mutations in this pathway were shown to generate MVP. Our study points to the importance of TGF β – LTBPs – fibrillin complex in normal valve physiology and its relation to pathogenesis of MVP. This pathway may present an opportunity for pharmacological targeting to modify disease progression, as suggested by the study of Ng ^23^

Our data recapitulates finding by others ^38–40^ by demonstrating lens dislocation and reduced visual acuity. The reduction in visual acuity observed in *LTBP2* KO mice can probably be attributed to the dislocation of the lens, a condition known as ectopia lentis. This parallels the human clinical manifestation of ectopia lentis, when progressive subluxation or complete dislocation of the lens can cause a high degree of myopia and reduction in visual acuity varies with the type and degree of dislocation. The lens dislocation in *LTBP2* KO mice likely disrupts the normal refraction of light onto the retina, impairing the sharpness and clarity of the visual image. Further investigation into the structural and functional integrity of the ocular components in *LTBP2* KO mice could elucidate the mechanistic pathways linking LTBP2 deficiency to lens stability and visual acuity.

Our study has several limitations: As of the writing of this article we have found only one family carrying the current mutation. However, the recent GWAS cited above that found a hit adjacent to the LTBP2 gene ^25^ provides confirmation from another cohort. Antonutti et al described that LTBP2 mutations are associated with spontaneous coronary dissection ^41^. Morlino has described Marfan like features in Roma/Gypsy subjects with the LTBP2 homozygous p.R299X variant ^42^. The latter two further support LTBP2 role in normal ECM stability in various tissues and that the phenotype may expand to other organs. The exact role of LTBP2 in the pathogenesis of MVP remains unclear. Future work will seek to identify and explore the specific molecular mechanisms that link LTBP2 and ECM homeostasis signaling pathways. We hypothesize that LTBP2 has a lifelong role in homeostasis and maintenance of the elasticity of the tissues.

### In conclusion

A recent large GWAS found an association between LTBP2 and MVP ^25^. We provide data on a pedigree with LTBP2 mutation linked to the trait. Our animal model data provides evidence for the important role of LTBP2 in normal mitral function and structure. Mutations in this gene cause myxomatous valve degeneration. This model may serve in the future for studies aimed at better understanding myxomatous degeneration pathogenesis and to test potential therapies.

## Data Availability

The authors confirm that the data supporting the findings of this study are available within the article [and/or] its supplementary materials.

## Non-standard Abbreviations and Acronyms

LTBP2: Latent Transforming Growth Factor Beta Binding Protein 2
MVP: Mitral Valve Prolapse
KO: Knockout
KI: Knock-in
WT: wild type mRNA messenger RNA
PCR: polymerase chain reaction q
PCR: Quantitative polymerase chain reaction
RUNX2: Runx family transcription factor 2
ECM: extra-cellular matrix gRNA guide RNA
CRISPR: Clustered Regularly Interspaced Short Palindromic Repeats sgRNA single guide RNA
SSC: Sequence scan for crispr
ssODN: single-stranded oligodeoxynucleotides
tracrRNA: trans-activating CRISPR RNA
crRNA: CRISPR RNAs
IDT: Integrated DNA Technologies
H&E: Hematoxylin and eosin staining
DAPI: 4’,6-diamidino-2-phenylindole
qRT-PCR: Real-Time Quantitative Reverse Transcription PCR
CTGF: connective tissue growth factor
GAPDH: glyceraldehyde-3-phosphate dehydrogenase protein family
MR: mitral regurgitation
GWAS: genome-wide association study
LTBPs: Latent Transforming Growth Factor Beta Binding Proteins
MFS: Marfan syndrome
FBN1: Fibrillin1
LAP: latency-associated peptide
OCT: Optical coherence tomography

## Acknowledgment

We thank Dr. Gerhad Sengle for providing LTBP2 antibodies for valve immunostaining. The paper was supported by BSF grant 2017265, and Hadassah Medical Center bridge grant in 2021. The authors report nothing to disclose related to the current publication.

---

**Supplementary S1:**
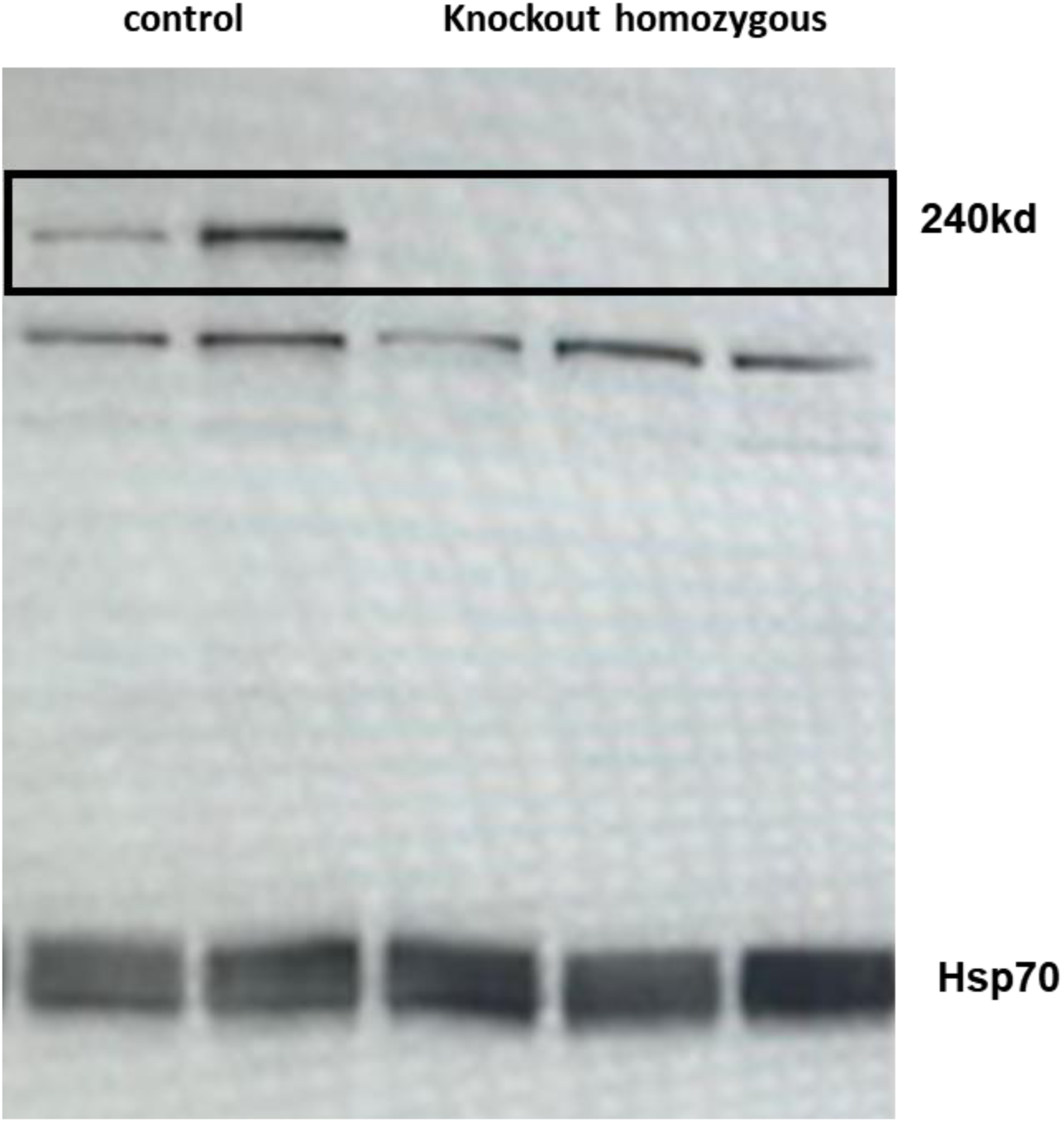
Western blot analysis of total proteins extracted from hearts of homozygous knockout mice versus hearts of control samples. Please note the bank size for LTBP2 is approx. 240 KD. A clear band is noted in the control animals while it is lacking in the KO animals.

**S2:**
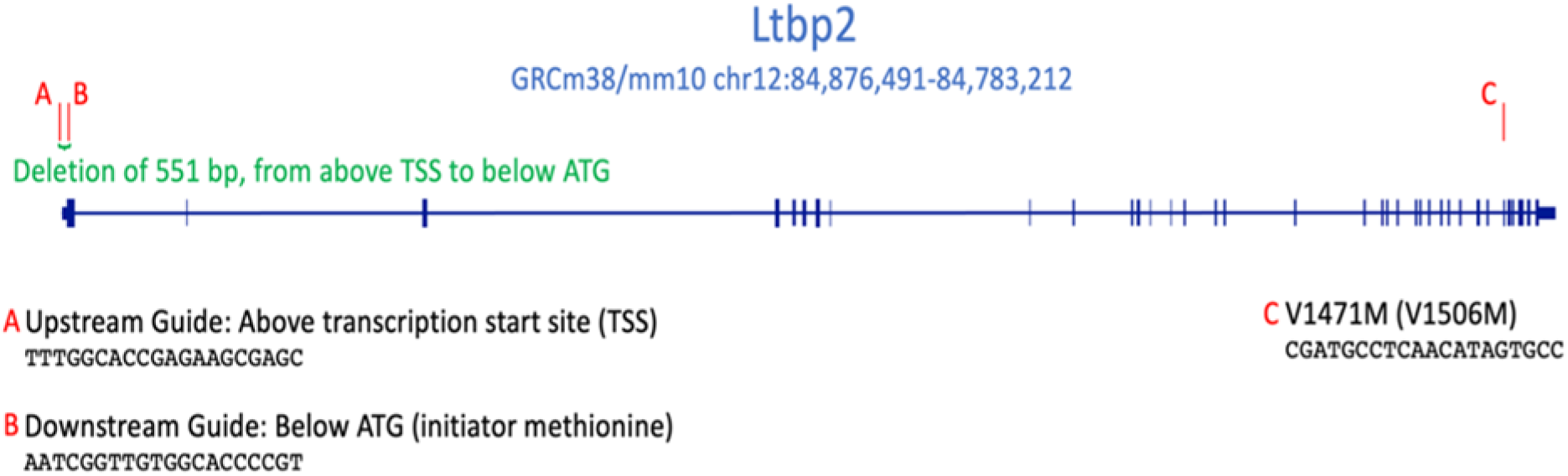
LTBP2 design scheme.

